# Adaptive Geospatial Surveillance System for Antimalarial Drug Resistance

**DOI:** 10.1101/2025.05.15.25327572

**Authors:** Apoorv Gupta, Lucinda E. Harrison, Minu Nain, Sauman Singh-Phulgenda, Rutuja Chhajed, Roopal S kumar, Aishika Das, Manju Rahi, Philippe J. Guerin, Anup R Anvikar, Mehul Dhorda, Jennifer A. Flegg, Praveen K. Bharti

**Author notes:** Corresponding authors: Dr. Praveen K. Bharti, Prof. Jennifer A. Flegg. Contributed equally (Apoorv Gupta, Lucinda E. Harrison). Contributed equally (Jennifer A. Flegg, Praveen K. Bharti).

## Abstract

Disease surveillance activities are usually resource-constrained and should be optimised to achieve spatial and real-time situational awareness. Such optimisation would help with better resource allocation, reduced logistics, and other costs. India has a high population density, diverse geography and climatic conditions, and difficult terrain. With respect to malaria, *Plasmodium falciparum* (Pf) and *Plasmodium vivax* (Pv) are endemic, with substantial variability of transmission across the country. While for Pv, drug efficacy appears to be homogenous within the country, for Pf malaria, the resistance pattern varies from the northeastern region to the central region. These factors make accurate mapping of antimalarial drug resistance difficult. To account for these complexities, we develop a targeted and adaptive methodology to identify prospective study sites for Pf antimalarial drug resistance surveillance. We retrieve existing data on the prevalence of validated markers of resistance to Artesunate (AS) and Sulfadoxine-Pyrimethamine (SP) from the World Wide Antimalarial Resistance Network (WWARN) systematic review database. We incorporate these data into a geostatistical model to estimate the prevalence of these markers across India and identify areas with high projected median resistance marker prevalence and low uncertainty. Finally, we create an interactive dashboard using the RShiny software package to simplify the process of selecting sites for future molecular surveillance. This methodology helps to ensure that decision-making is supported by data and modelling outputs while facilitating the generation of knowledge about the current state of antimalarial drug resistance with wide geographic coverage. We demonstrate the utility of our method by selecting sites for surveillance of drug resistance in Pf malaria in India.

## Introduction

Malaria remains a significant public health problem despite global efforts to control and eliminate it (World Health Organization, 2021)(1). The World Health Organization (WHO) in its Global Technical Strategy (GTS), adopted in May 2015, has set a goal of 90% reduction in malaria cases by 2030 (World Health Organization, 2015)(2). Antimalarial drug resistance surveillance aims to identify, monitor, and map regions where drug-resistant malaria strains are prevalent, to allow for effective treatment strategies to be put in place(3-6). Antimalarial drug resistance surveillance involves the collection and analysis of data on malaria transmission, drug efficacy, and patient outcomes (World Health Organization, 2021)(1, 4). Surveillance data may then be used to inform public health policies and interventions, such as the deployment of alternative antimalarial drugs or diagnostics, as well as the implementation of vector control measures to prevent malaria^4^.

Surveillance is a resource-intensive and time-consuming process. Available resources must be utilised optimally to improve surveillance coverage and achieve close to real-time situational awareness, ultimately leading to better resource allocation. Recent work has shown that mathematical models can provide additional resources for improving disease surveillance as they allow for predicting and visualising disease transmission patterns, treatment efficacy, and the impact of intervention strategies(7, 8). Geospatial models can characterise the spatial distribution of malaria cases and identify high-risk areas for targeted intervention(7). Geospatial modelling has been used to support antimalarial resistance surveillance efforts by identifying areas of highest value to surveillance, given both predicted resistance prevalence and uncertainty in model predictions(9, 10).

Following the recommendation of the WHO GTS, India launched its ‘National Framework for Malaria Elimination’ in 2016. Under this framework, India aims to eliminate malaria, prevent reintroduction, and maintain malaria-free status across the country by 2030 (Ministry of Health and Family Welfare, 2016). With the national increase of *Plasmodium falciparum* (Pf) malaria cases, the rise of AS+ SP resistance, which is a commonly used drug to treat Pf malaria, and instances of K13 resistance in the north-eastern region, mapping of antimalarial resistance will become a priority to achieve the goals of the National Framework for Malaria elimination by 2030(11, 12). Surveillance of validated resistance gene mutations present in malaria parasites is common practice to capture the emergence of antimalarial drug resistance.

India has complex geographical boundaries, varying temperatures and climates, and vast river basins Pf and Pv are endemic across the country, with large heterogeneity in prevalence and transmission. Approximately 130,000 Pf malaria cases are reported annually, causing the death of 82 patients. The current recommendations of uncomplicated Pf malaria are AL in the northeast region of India and AS-SP in the rest of India because of occurrence of (AS-SP) resistance in northeast region. Different regions in India require different Pf malaria drug interventions, adding to the complexity of surveillance efforts (Ministry of Health and Family Welfare, 2016). To optimise current surveillance efforts, a targeted methodology is needed to identify prospective study sites where antimalarial drug susceptibility data are urgently needed(13). The use of geospatial modelling tools and dynamic dashboards can aid surveillance efforts by identifying priority areas based on specific decision objectives and constraints(14, 15).

We develop a methodology for informed antimalarial drug resistance surveillance using tailor-made geospatial models for India. The models use existing prevalence data from India on genetic markers of antimalarial drug resistance, including mutations in *P. falciparum* dihydropteroate synthase (*pfdhps*), dihydrofolate reductase (*pfdhfr*) and in *pfk13*. Source data is retrieved from the World Wide Antimalarial Resistance Network (WWARN) molecular systematic review database and supplemented by published data identified from a recent systematic review(16). We use these data to inform geostatistical models of resistance marker prevalence, which provide both a surface of predicted marker prevalence for India and a surface of prediction uncertainty. To facilitate site selection, we create a dashboard using the RShiny software package(17). The districts selected using the RShiny dashboard are presented in visual and tabular formats with the districts ranked based on a set of epidemiological parameters. The dashboard facilitates data-driven decision-making and visualises the current status of antimalarial drug resistance in India. Our methodology is piloted to identify sites for future molecular surveillance.

## Methodology

### Data Extraction from WWARN DHPS and K13 systematic review database

In this study, we used data on the prevalence of *pfdhfr/pfdhps* and *pfk13* mutations across India. The data was obtained from previously established databases of *pfdhps/pfdhfr* and *pfk13* resistance markers accessible on the WWARN molecular surveyors (https://www.iddo.org/wwarn/tracking-resistance). The WWARN surveyor has been recently updated with the data extracted from published and unpublished studies on the molecular prevalence of *P. falciparum* markers of resistance to SP (*pfdhps, pfdhfr*) and to artemisinin (*pfk13*) across India up to March 2022. In addition, we conducted a systematic literature review to identify additional data from India. We followed the identical methodology similar to the Minu Nain et al.(16) study that searched strategy, PRISMA flowchart, inclusion and exclusion criteria, etc.(9) The updated information on the WWARN SP and the artemisinin resistance surveyors were then used for the geospatial modelling and identification of surveillance sites. Data were extracted only from those studies in the WWARN database that reported complete information on the location and period of sample collection, basic demographic data, and the prevalence of the markers of interest. Extracted data contains study title, author details, study year (precise or estimated), publication year, geographical location (precise or estimated), age group, gender, number of successfully sequenced samples for each study site, number of samples with wild-type parasites and with WHO-validated single nucleotide polymorphisms (SNPs) associated with SP resistance (*pfdhps*/*pfdhfr*) or artemisinin resistance (*pfk13*) were recorded. Strains carrying the *pfdhps540E* mutation are uncommon, and the mutation is typically seen in conjunction with additional *pfdhps* and *pfdhfr* mutations in the form of quintuple and sextuple mutations. As a result, *pfdhps540E* is commonly utilised as a proxy for quintuple and sextuple PfDHFR/PfDHPS mutations linked with clinical SP resistance.

### Spatiotemporal statistical modelling

Predictive maps of *pfdhps540E* and the relevant *pfk13* mutation prevalence were created using the data extracted WWARN Surveyors(18). The *pfdhps540E* and *pfk13* datasets were used in a Bayesian geostatistical model to produce predictive maps of the prevalence of the markers in India. The statistical methodology for spatiotemporal prediction has been documented previously and follows two stages(10, 19, 20). In the first stage, the posterior distribution of model parameters given the observed data was estimated. Convergence diagnostics for the Bayesian inference were monitored, including trace plots, autocorrelation and coverage probabilities; no evidence of divergence was found. In the second stage, the posterior predictive distribution of marker prevalence was predicted on a 5 x 5 km^2^ resolution over India, given the posteriors for the model parameters from the first stage. The posterior predictive distribution at each location was summarised using the median and standard deviation to create point estimates and uncertainty surfaces, respectively. We consider modelling outputs relevant to 2021-22 in the remainder of the workflow, as this is the year for which other epidemiological data is available. We visualise the model outputs using Administrative India Boundary shapefiles up to district level with HQ-Scale 1:1M (having State Boundary and District Boundary) from survey of India with product code OVSF/1M/7.

### Epidemiological datasets employed

In this study, we consider three malaria-specific epidemiology variables for categorising states and their subsequent districts, source: (Malaria Annual Report 2023, from National Center for Vector Borne Disease Control (NCVBDC), Ministry of Health and Family Welfare, Govt. of India):

1. Total number of Blood Slide Examination (BSE) - Total number of blood slides examined under a microscope in a month to detect malaria parasite infection.
2. *Plasmodium falciparum* (Pf)-Total number of Pf malaria-positive cases per month with positive blood slide examination.
3. *Plasmodium falciparum Parasite Rate* (PFPR) - Total rate of *Pf* malaria positivity in a month:

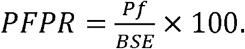

These parameters are integrated into the dashboard for ranking and clustering states and their subsequent districts into three groups of mild, moderate, and high Pf malaria burden.

### R Shiny Dashboard

An RShiny dashboard was constructed to use modelling outputs and epidemiological data to facilitate the selection of districts for surveillance(21). The dashboard allows the user to place a set of “filters” in a preferred order to exclude districts on the grounds they do not satisfy one of the filtering criteria. For example, a “filter” can be placed on the Pf parasite rate (PFPR), to include only the 50 districts with the highest PFPR in further decision-making. The dashboard allows the results of multiple filtering outputs to be visualised and compared without the decision-maker being familiar with the R programming language or any of R’s functionality for geospatial modelling or visualisation. The workflow featured in our dashboard was iterated subject to project aims and data availability – the final workflow excludes districts based on filtering variables (i.e., a subtractive approach), rather than optimising one or more decision objectives (a more direct approach). The set of districts selected through the dashboard is highly sensitive to decisions made during the selection and ordering of filters, and the number of districts excluded as each filter is applied. This sensitivity is acknowledged, but allowable: the workflow presented is a quantitative, structured approach to decision-making and is preferable to selecting sites ignoring available data and without considering specific surveillance objectives (e.g., selecting equidistant sites or selecting sites at random)(22, 23). A version of the dashboard featuring published data is available at https://lucyharrison.shinyapps.io/pf_drug_resistance_shiny/.

### Workflow for site selection

We selected four districts for surveillance, based on the geospatial modelling of resistance markers and other epidemiological data across India (Figure 1). In the initial step, geolocated data of resistance marker prevalence in India and adjacent areas were extracted from updated WWARN K13 and DHPS/DHFR molecular surveyors. Using these data, two geospatial models were constructed as described in the spatiotemporal statistical modelling methodology section. Following model validation, both models were integrated into the Rshiny dashboard with 14 other epidemiological variables for filtering and ranking districts to identify sites for prospective surveillance. The workflow for district selection was agreed on after multiple iterations of possible decision objectives and constraints. The final workflow divided districts into three clusters of malaria endemicity (high, medium, and low). Pragmatic considerations including site feasibility, population density, and expected number of malaria cases, were used to finalise site selection from the filtered district-level outputs.

**Figure 1:**
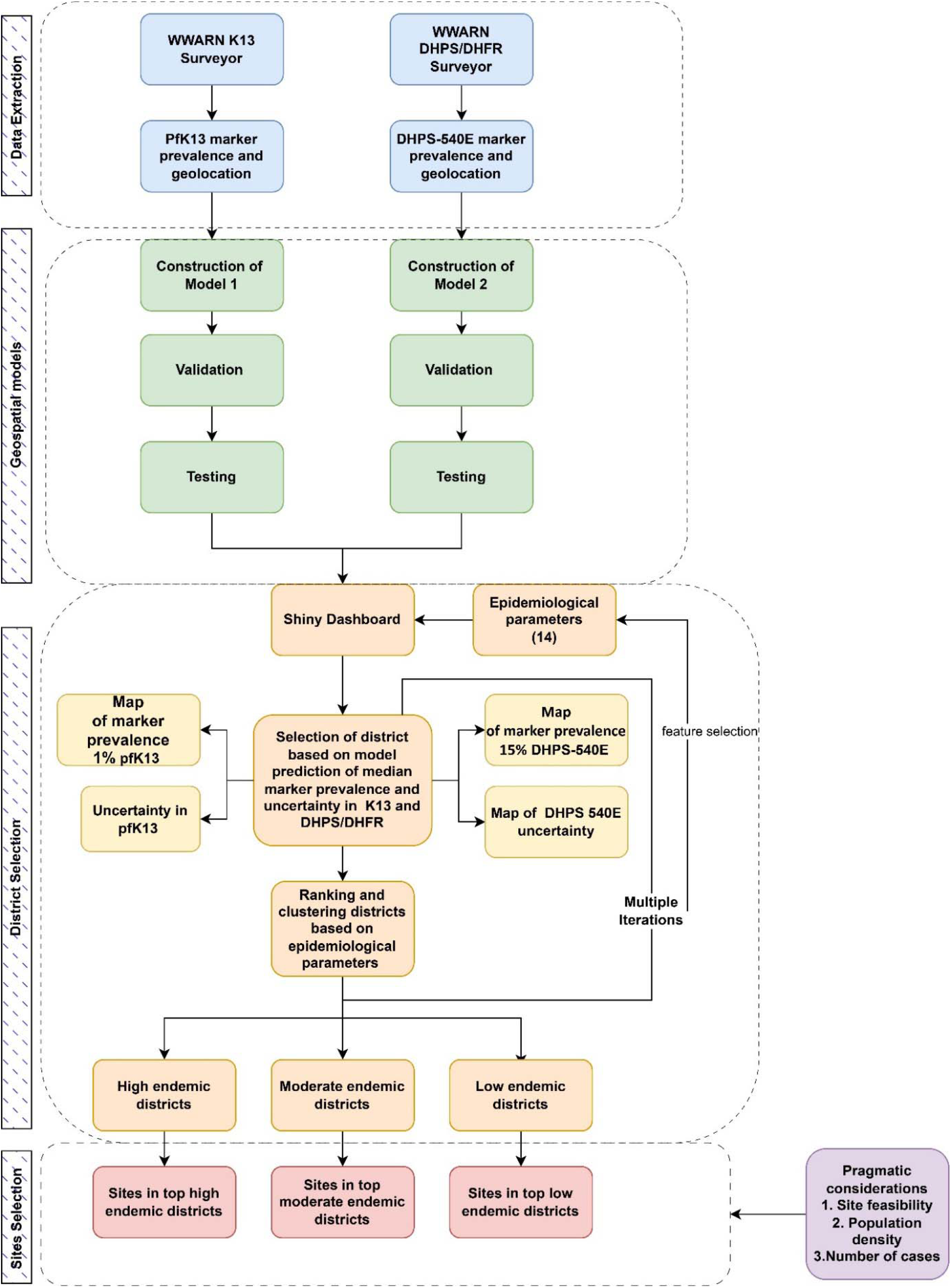
Flowchart describing the modelling and decision-making workflow: data are extracted to inform geospatial models; the outputs of geospatial models, together with other epidemiological parameters, inform district selection; surveillance sites are selected from filtered districts.

## Results

### Data summary

The extracted dataset from WWARN Surveyors contains 113 *pfk13* and 67 *pfdhps*540E resistance marker records in India, reported between 1994 and 2019. We used the *pfdhps*540E geospatial model (with zero-time separation and a spatial correlation threshold of 1%) to calculate a reasonable cut-off distance of 675 km from the Indian border. This included studies that could affect the model from neighbouring countries; a total of 54 *pfk13* studies and 40 *pfdhps* studies having 155 *pfk13* data entries and 24 *pfdhps*540E entries, respectively, were extracted from six neighbouring countries, including Afghanistan, Pakistan, Nepal, Bangladesh, China, and Myanmar. Most of the Indian studies for *pfdhps*540E are from the East and Northeast regions. There are also 15 entries from the Central regions of India and 6 entries from the North for *pfdhps*540E. Correspondingly, most entries in the *pfk13 d*ataset are from East and Northeast regions, 20 from Central, and a minority from the North (n=4), South (n=4), and West (n=2) regions of India.

### Geostatistical model

Using the data, continuous predictive maps for *pfk13* and *pfdhps*540E from 2000 to 2022 were generated. The construction of the statistical model allows predictions to be made at locations in space and time where no data is available, and the models allow for quantification of uncertainty in predictions.

The median of the posterior predictive distribution of *pfk13* was near zero over the entire country in 2021, except for a few ‘hotspots’ in the Northeastern region (Figure 2(a), Figure 2(c)). The associated uncertainty map (Figure 3(a)) showed relatively high certainty in these model results, but less certainty at the hotspots in the Northeast and Northwest regions. The median of the posterior predictive distribution of *pfdhps*540E was low across most of the country in 2021 but moderate in the Northeast, East, and Central regions (Figure 2(b), Figure 2(d)). The associated uncertainty is high in regions of moderate median values (Figure 3(b)).

**Figure 2:**
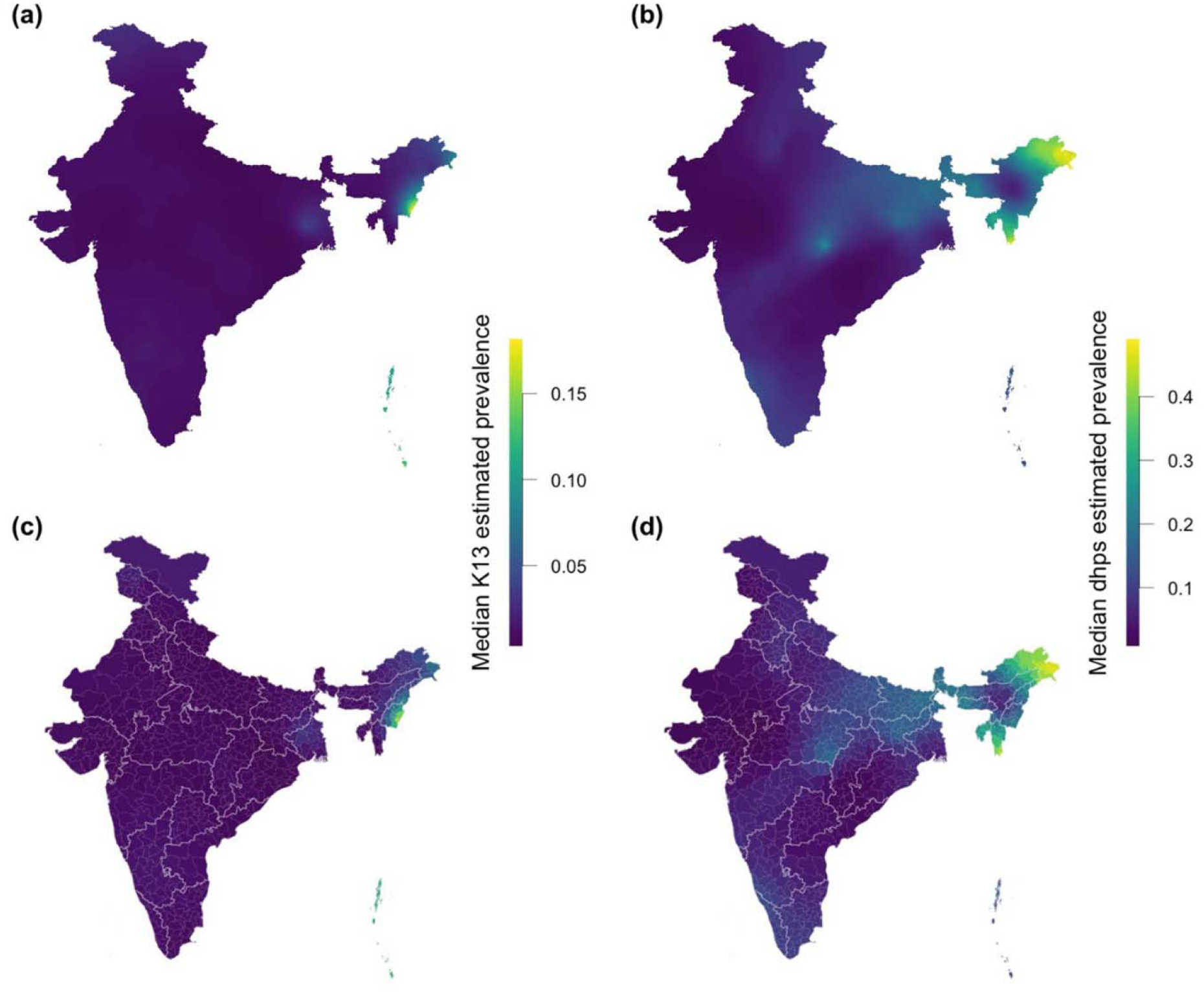
(a) Median posterior predictive prevalence of *pfk13* in 2021; (b) median posterior predictive prevalence of *pfdhps540E* in 2021; subplots (c) and (d) show the median posterior predictive prevalence by district for *pfk13* and *pfdhps540E*, respectively.

**Figure 3.**
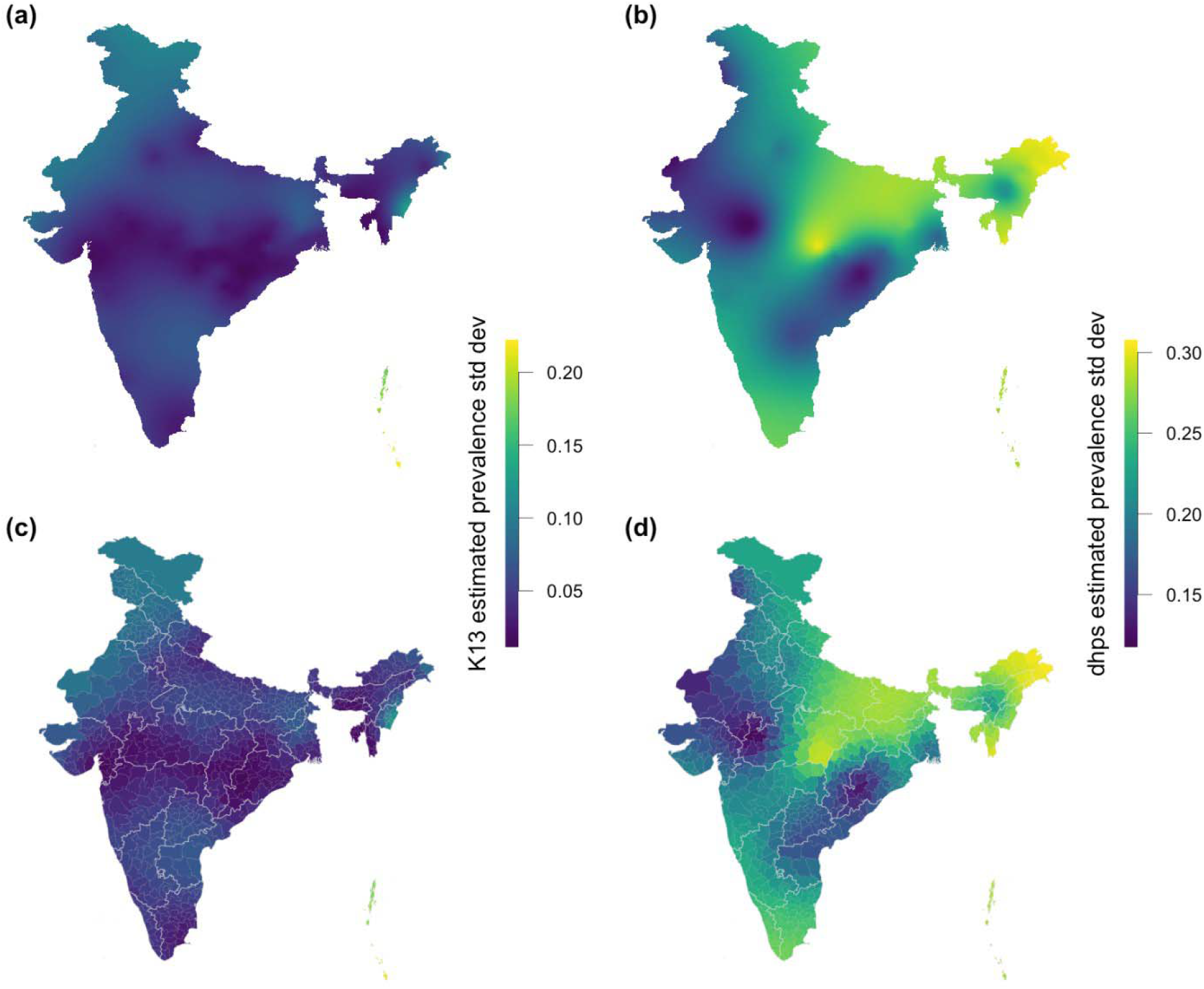
(a) Standard deviation for *pfk13* posterior predictions in 2022, associated with median predictions in Figure 2(a); (b) standard deviation for *pfdhps540E* posterior predictions in 2021, associated with median predictions in Figure 2(b). Subplots (c) and (d) show the standard deviation of posterior predictive prevalence by district for *pfk13* and *pfdhps540E*, respectively.

Out of 766 total districts across India, 200 were selected in the first step of the workflow as those with the highest *pfk13* prevalence median (Figure 4). Out of these 200 districts, 150 were selected as those having the highest *pfk13* prevalence standard deviation (SD) followed by the shortlisting of 100 districts with higher *pfdhps* median. Further, we ranked the 100 districts based on lowest to highest *pfdhps* SD in them. The latest 2022 state (PFPR) data was used to divide all states into the categories of high, medium, and low malaria burden – these categories were then assigned to the districts within each state. Of the shortlist of 100 districts, four districts were selected: one each from high and low-burden states and two from medium-burden states based on pragmatic considerations.

**Figure 4:**
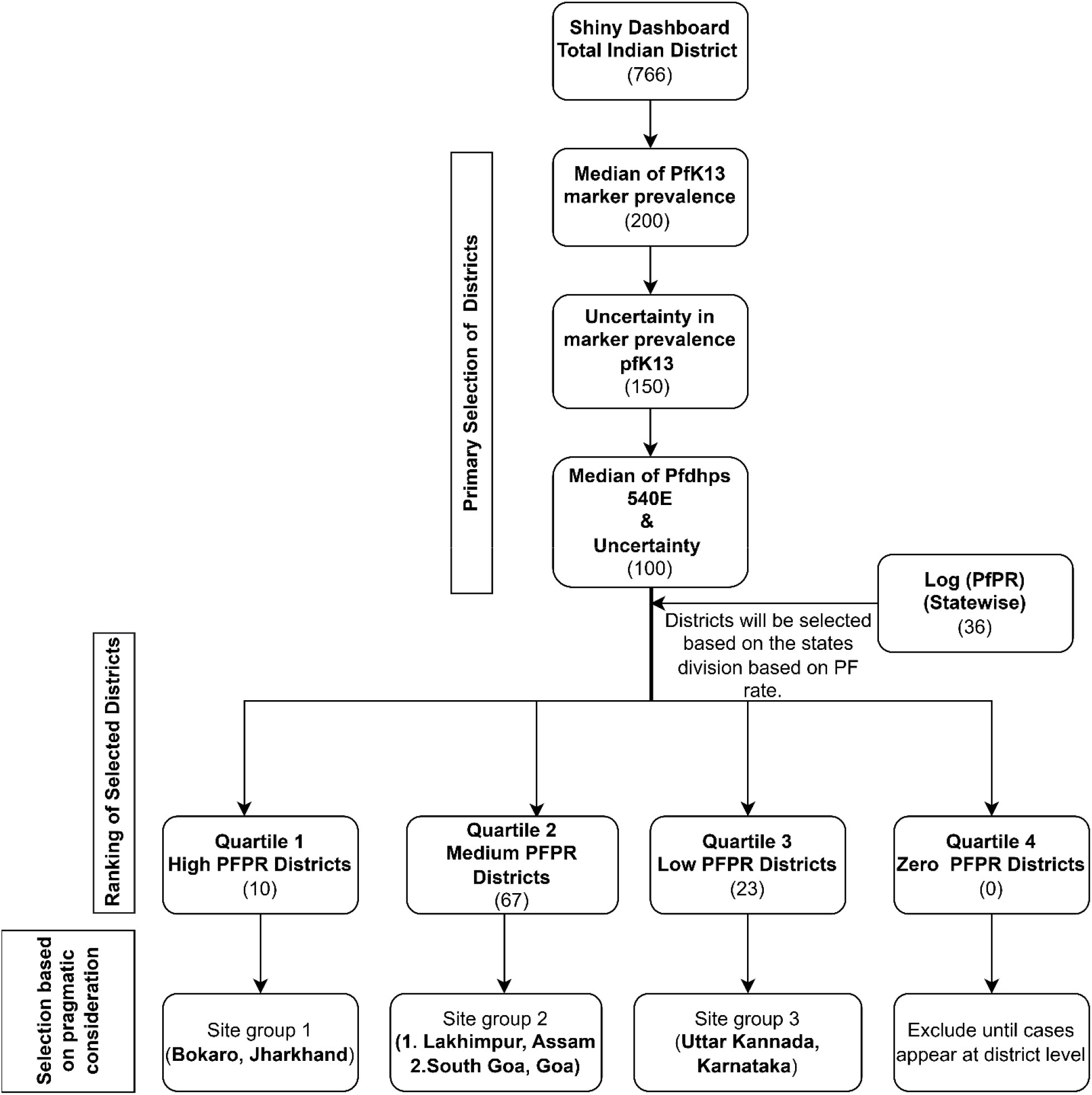
Flow diagram for district/site selection using the RShiny dashboard.

These are Bokaro in Jharkhand, Lakhimpur in Assam(24), South Goa in Goa, and Uttar Kannada in Karnataka(25) (Figure 5). These areas were selected based on available information on Pf malaria endemicity in the district or in adjacent areas, population density, and feasibility of surveys based on access, infrastructure, and local support etc.

**Figure 5:**
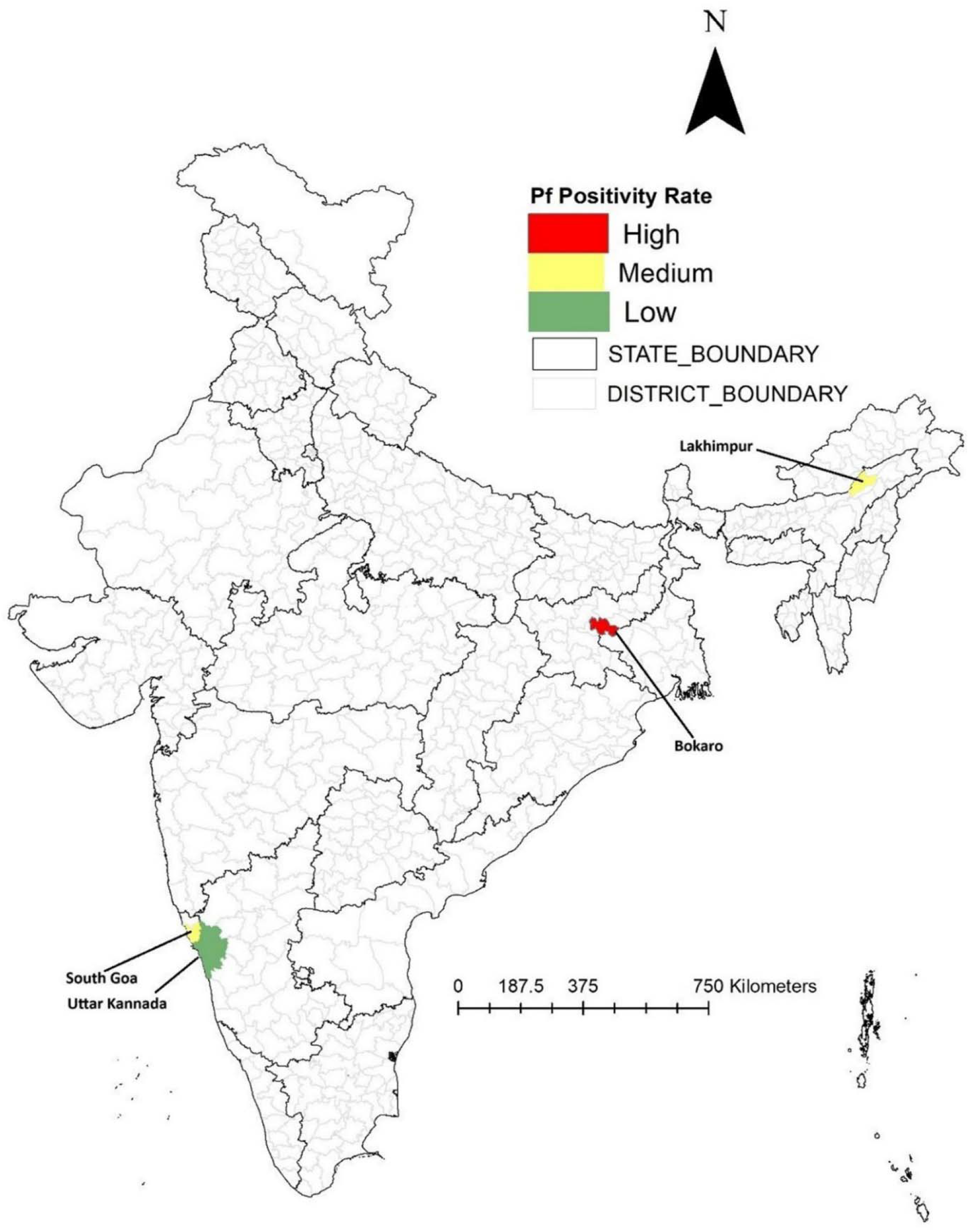
Location of four districts selected from geospatial modelling with colour scheme according to their PFPR.

## Discussion

In this paper, we describe a methodology to use geostatistical modelling of existing data to optimise surveillance of antimalarial resistance. Our workflow identifies areas of the highest value to surveillance, i.e., those with high predicted prevalence of parasite genetic markers of resistance and high uncertainty in model predictions, while also incorporating other relevant epidemiological parameters. The implementation of our modelling and decision-making workflow is facilitated by an RShiny dashboard app, that assists the systematic selection of surveillance sites to enhance surveillance coverage and increase the capacity for early detection of emerging resistance. We illustrate the utility of our workflow by piloting its implementation to select sites for molecular surveillance in India.

The prevalence of molecular markers indicating SP resistance in surveys conducted from 2001 to 2022 is elevated in the Eastern and Northeastern regions of India, as has already been confirmed from field surveys and modelling. Artemisinin resistance markers remained mostly limited to Northeastern states with an exception in West Bengal, related to a single study in which artemisinin resistance was identified; artemisinin resistance had not been detected in this region before and has not been detected since. The corresponding map of uncertainty in model estimates show high uncertainty in the Northeastern and Southern regions of the country. The results presented here on the distribution of genetic markers of antimalarial resistance include data from neighbouring countries within 675 kilometres of Indian borders, as these can influence the predictions of resistance prevalence made by our geostatistical models. Though this leads to slight variance from previously published results, our results however remain mostly similar.

It may appear paradoxical that the predicted estimates with the highest uncertainty are in regions where most surveys have been conducted. However, this is expected given that the geospatial model takes into consideration all results collected from a location or within a region, including those from before resistance emerged in those foci. As the presence or prevalence of resistance markers is confirmed over time and with multiple surveys in agreement, the uncertainty would be expected to decrease in these regions, e.g. as resistance reaches saturation.

The predicted prevalence and the associated uncertainty are key determinants to identify sites or districts where performing a survey would be most valuable, i.e. those which can confirm the presence or absence of resistance while also reducing the uncertainty in the modelled estimates. However, these surveillance objectives must be complemented by epidemiological, demographic, and other parameters to ensure that the surveys are feasible. Here, districts were first ranked with respect to the modelled prevalence of *pfk13* mutants, then the related uncertainty, followed by ranking based on the prevalence and uncertainty of *pfdhps* 540E. The shortlisted districts were then classified based on state-level PFPR, to ensure the malaria endemicity spectrum was covered in the final set of selected sites. This allows a balance between the collection of a reasonable number of specimens from malaria patients and enhancing coverage in areas with low endemicity where resistance may be more likely to emerge(26). The RShiny dashboard was developed to facilitate and systematise the iterative shortlisting of sites by integrating both geospatial model output and malaria epidemiology data. Finally, four districts, one each from states with high and low PFPR as well as two from states with moderate PFPR, were selected from the shortlist for this pilot exercise. This final selection was based on pragmatic considerations – population density, historical malaria case burden, and existing human resources and/or infrastructure to facilitate survey implementation. Prospective surveillance is being conducted at all these sites and the prevalence results will be used for a second round of geospatial modelling along with any other newly generated or existing data.

The estimated prevalence determined from the modelling work presented here may or may not be confirmed by the ongoing field surveys at the selected sites. Resistance marker prevalence data is scarce in the study area, contributing to model uncertainty. There was very limited access to the recent district-level malaria endemicity data for site filtering - this would provide more spatial precision than state-level data. It is also possible that there are other factors, such as human mobility or access to treatments (i.e., selective pressure on the parasites), which can influence the spread or emergence of resistance but which are not yet incorporated in the geospatial model. Depending on the results obtained from initial rounds of surveys, the model may need to be refined and/or augmented to include additional parameters to improve its accuracy. However, in its current form, it allows a consistent approach to systematic surveillance for antimalarial drug resistance.

## Conclusion

Effective disease surveillance requires optimal use of limited resources and an understanding of the target phenomenon’s spatial distribution. The methodology described in this paper provides a systematic way to optimise surveillance for antimalarial drug resistance while improving the estimates of its distribution across India and its diverse demographic, topographic, and climatic contexts. The use of a dynamic dashboard created with the RShiny software package ensures decision-making is quantitatively supported by modelling outputs while keeping stakeholders informed about the current state of antimalarial drug resistance. Our methodology will be particularly useful in contexts with scarce data to optimise and target surveillance efforts as they are scaled up.

## Data Availability

The source data used for modelling is available from Artemisinin Molecular Surveyor (http://www.wwarn.org/molecular/surveyor/k13/index.html?t=201608031200#0) and SP Molecular Surveyor (http://www.wwarn.org/dhfr-dhps-surveyor/#0).
R code is available on GitHub and can be access through below link (https://github.com/lu-harr/pf_drug_resistance). At present the dashboard has not been uploaded online due to data protection issues but a proof of concept on simulated data can be accessed through https://lucyharrison.shinyapps.io/pf_drug_resistance_shiny/.

http://www.wwarn.org/molecular/surveyor/k13/index.html?t=201608031200#0

http://www.wwarn.org/dhfr-dhps-surveyor/#0

## Declarations

### Author contributions

All authors were involved in the conception and design of the study and reviewed all related documents and materials. AG and MN did data extraction from WWARN repository, and AG and LEH wrote the first draft of the manuscript. RSK and AD assisted in manuscript writing. JF performed geospatial modelling. LEH and AG assisted JF in geospatial modelling and preparation of figures. LEH developed the shiny dashboard and AG developed the final pipeline using dashboard for sites identification. MD coordinated the study, designed the protocol, reviewed and provided inputs on the methodology and manuscript. AG also prepared figures 1, 2, and 5. JF ran geospatial models and prepared figures 4, 5, and 6. SSP and RC assisted with the coordination of the study. PB and MR supervised the study activities. PJG and ARA conceived the idea and reviewed the manuscript. All authors read and approved the final manuscript. AG, LEH and JF are the guarantors for this work.

### Data sharing Statement

The source data used for modelling is available from Artemisinin Molecular Surveyor (http://www.wwarn.org/molecular/surveyor/k13/index.html?t=201608031200#0) and SP Molecular Surveyor (http://www.wwarn.org/dhfr-dhps-surveyor/#0).

R code is available on GitHub and can be access through below link (https://github.com/lu-harr/pf_drug_resistance). At present the dashboard has not been uploaded online due to data protection issues but a proof of concept on simulated data can be accessed through https://lucyharrison.shinyapps.io/pf_drug_resistance_shiny/.

